# Estimation of Case Fatality Rate during an Epidemic: an Example from COVID-19 Pandemic

**DOI:** 10.1101/2020.05.19.20106468

**Authors:** Masud Yunesian, Akbar Fotouhi, Mina Aghaei

**Affiliations:** Department of Environmental Health Engineering, School of Public Health, Tehran University of Medical Sciences, Tehran, Iran; Department of Research Methodology and Data Analysis, Institute for Environmental Research, Tehran University of Medical Sciences, Tehran, Iran.; Department of Epidemiology and Biostatistics, School of Public Health, Tehran University of Medical Sciences, Tehran, Iran

## Abstract

The case fatality rate (CFR) is an important index in epidemics and policymakers need to be aware of this statistic for making sound decisions. However, there are some uncertainties in the calculation of the fatality rate and the true case fatality rate can be calculated after an epidemic subsides, we calculated the fatality rate for 20 countries which had the highest number of confirmed cases with three scenarios. In this study, we provided detailed information to discuss the studies that calculate the case fatality rate during the COVID-19 pandemic and highlight the estimation bias in these studies.

---

More than 2027000 patients tested positive by April 15^th^ around the world and 129104 deaths have occurred among these people resulting in a gross case fatality rate of 6.36 percent. The case fatality rate (CFR) is an important index in epidemics and policymakers need to be aware of this statistic for making sound decisions. There are, however, some uncertainties in the calculation of the fatality rate. The true case fatality rate can be calculated after an epidemic subsides, as the number of deaths will be from the total number of people who had contracted the disease after the subsidence of epidemics. We, however, may need to calculate this index while combatting the epidemics. One way is to go back for a certain amount of time for enumeration of the denominator of the case fatality rate (the total number of patients who became sick at the same time as those who died). There is not, however, consensus on how much we should go back to reach the true denominator. This is somewhat related to the stage of the disease that patients are being diagnosed. The more complete the case finding strategy, the earlier we diagnose the patients and the longer the time between diagnosis and death.

Manuel et al. produced three scenarios with 2, 4 and 7 days lag time from diagnosis to outcome determination and showed a wide range of uncertainty of fatality rate in these scenarios (1). Some believe that we should go back even more and use the number of confirmed cases of two weeks earlier as the denominator of the case fatality rate.

Hung C. et al. and Wang D. et al assumed that the median time from onset of symptoms to admission on intensive care unit would be around 10 days, whereas WHO announced that there may be 2 to 8 weeks interval between onset of symptoms and death (2-4). So, some researchers suggested dividing the cumulative number of cases in each day by the cumulative number of confirmed cases of 14 days earlier.

Baud et al. produced a graph comparing the apparent fatality rate used by WHO with fatality rate calculated with 14 days delay and concluded that the latter may be more accurate (5).

On the other hand, routine statistics of “confirmed cases” usually include more severe cases and true cases may be much more than the reported ones on a magnitude of several times.

The situation will be even more complicated if we are interested in “infection fatality rate” as the latter focuses on the total number of infected people regardless of their symptomatic situation. In fact, case fatality rate is always greater than infection fatality rate in diseases with asymptomatic infections, though this distinction seems not to be the case in Covid-19 as the majority of infections are believed to become symptomatic. In countries where diagnostic tests are reserved for the most advanced cases, the majority of mild to moderate cases will not be enumerated. There are two factors that act in different directions: the first one tends to underestimate true case fatality rate (as we include all diagnosed cases up to the time of calculation), and the second one will bias the true rate upward due to failure to include mild to moderate cases in the denominator. The results of these factors will depend on the relative magnitude of each one. We believe that the second factor is possibly much bigger generally, so the true case fatality rate could be much less than the number that is calculated routinely. The delayed calculation method is misleading in some situations especially in the first days of the epidemics. The first cases are very close (or even equal) to the number of deaths. In fact, in the early stages of the epidemic, only severe or fatal cases may be noticed by health authorities. Thus, the case fatality rates at the beginning of the epidemics in Wuhan were much bigger than the true fatality rate(s), as can be seen in the graph prepared by Baud *et.al (5)*.

We calculated the fatality rate for 20 countries which had the highest number of confirmed cases up to April 15^th^ with three scenarios. The first scenario assumes routine WHO figures (enumeration of cases and deaths on the same day). One-week and two-week delays were assumed for the second and third scenarios respectively. As is shown in Fig. 1, apparent fatality ranges from 0.8% (Russia) to greater than 13.2% (Belgium) with a global value of 6.8%.

**Figure 1.**
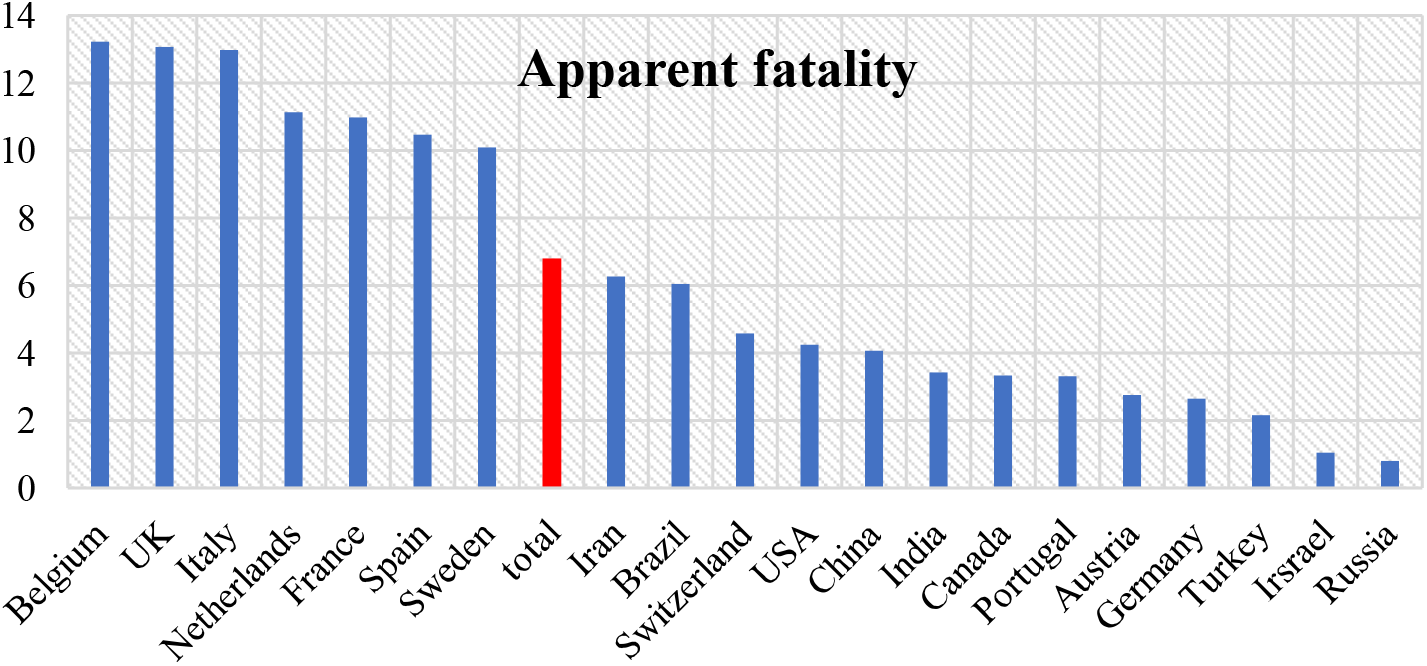
The apparent fatality rate for 20 countries with the highest number of confirmed cases of COVID-19 up to April 15^th^.

If we use the one-week delay calculation (Fig. 2), fatality rates will increase by about 33% globally (from 6.8% to 9.04%) and ranges from 1.33% (Israel) to 21.18% (UK). These values will increase more in the third scenario (two-week delay) ranging from 2.07% (Israel) to 43.65% (UK) and the global fatality will be around 14.73% (Fig. 3). In fact, UK, Belgium, and France will produce fatality rates greater than 30% in the third scenario. As the epidemic progresses, if we repeat this calculation, we will notice that the results are be very high in the exponential phase of epidemic and will decrease by the start of plateau phase. So, it seems not reasonable to assume a constant delay between diagnosis and death, especially when the number of cases increases exponentially and the majority of diagnoses (and confirmed) cases are very severe ones. We suggest, instead to focus on countries that are in the plateau or descending phase of epidemics e.g. China. As is seen in these figures, the fatality rate remains pretty constant in all scenarios for China (4.06%, 4.08%, and 4.09%) respectively. The best estimates, however, will be achieved if we select a cohort of patients whom we are aware of their ultimate situation (recovery or death).

**Figure 2.**
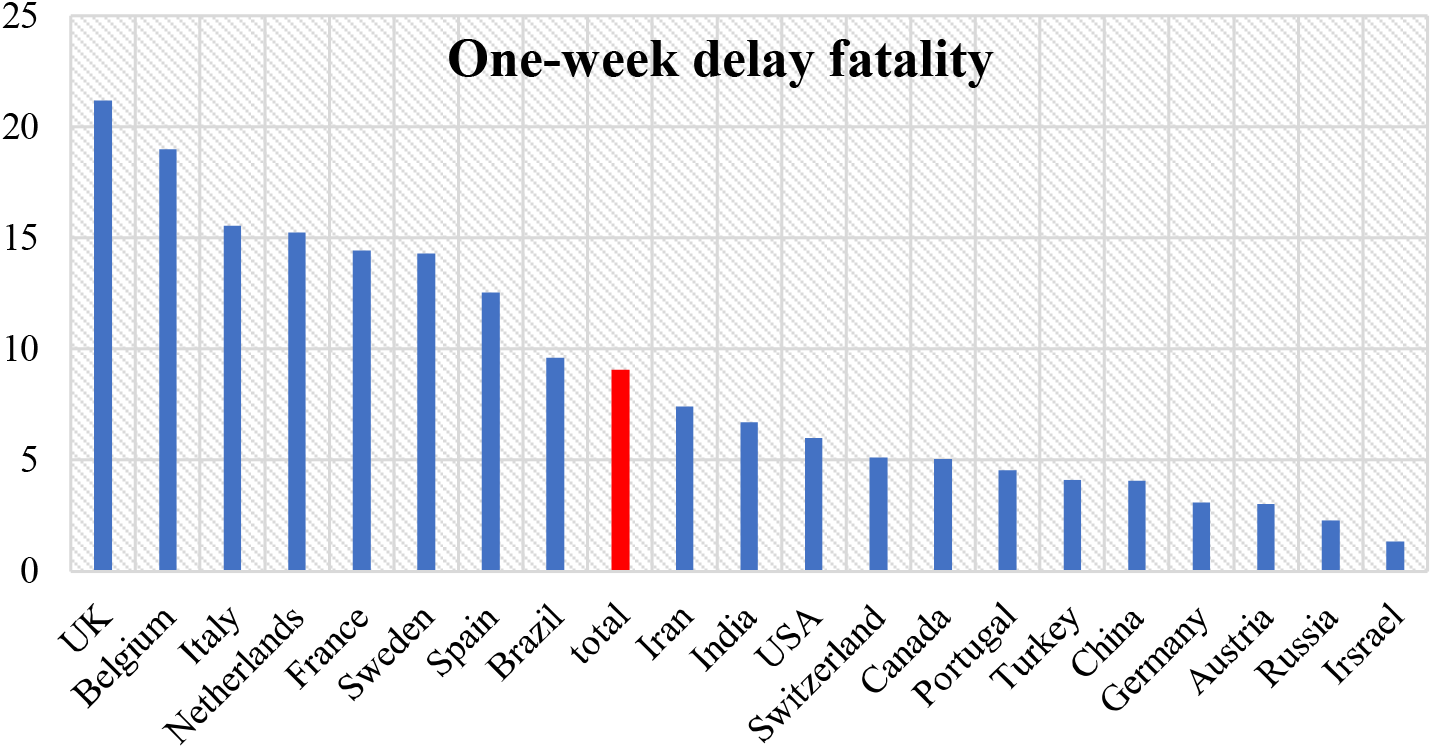
The one-week delay fatality rate for 20 countries with the highest number of confirmed cases of COVID-19 up to April 15^th^.

**Figure 3.**
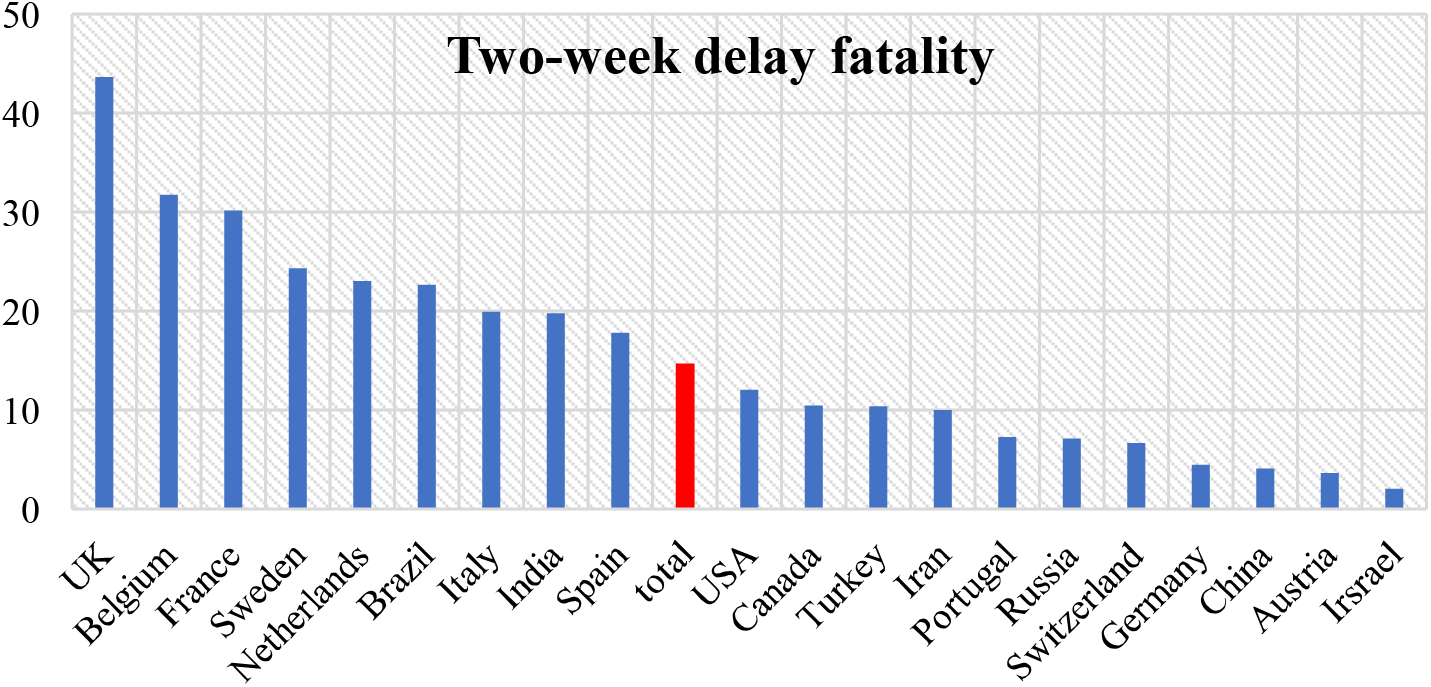
The two-week delay fatality rate for 20 countries with the highest number of confirmed cases of COVID-19 up to April 15^th^.

Since days from the first symptom to death are different in countries (ranges from 6 to 41 days) (6), one can approach alternative method to estimate the CFR during an outbreak by dividing the cumulative number of deaths by the sum of total recovered cases and cumulative deaths at any given time points as follows:

***[CFR = deaths / (deaths + recovered)]***.

Figure 4 shows the results of this estimation with a global value of 21.5%. However, as average length of stay of patients in hospital vary from country to country, estimation of the case fatality rate using this model is not precise too. This approach assumes that risk of death among active cases will be the same as the sum of deaths plus recovered. However, we know that risk of mortality among active cases will be much lower than the latter, as the majority of critically ill patients have died so far.

**Figure 4.**
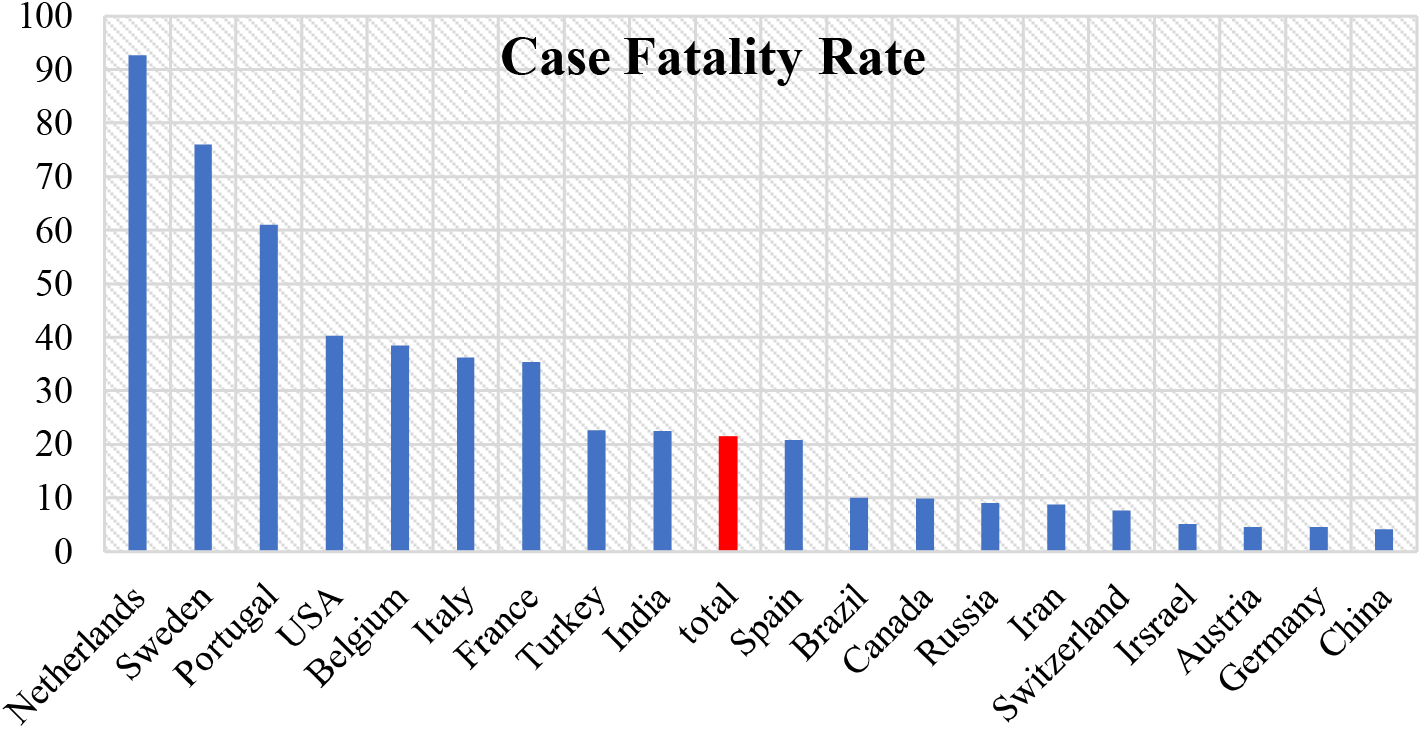
The fatality rate for 20 countries with the highest number of confirmed cases of COVID-19 using alternative model – (data up to April 15^th^)

Finally, we conclude that although apparent fatality rate increases and delayed fatality rates decrease by the progression of epidemics, ***using fixed delayed scenarios*** for calculation of case fatality rate may lead to ***serious bias*** (misleading) and should not be used. The third approach (deaths / (deaths + recovered)) will also bias the results upward and none of these approaches can produce unbiased estimate of case fatality rate (Figure 5).

**Figure 5.**
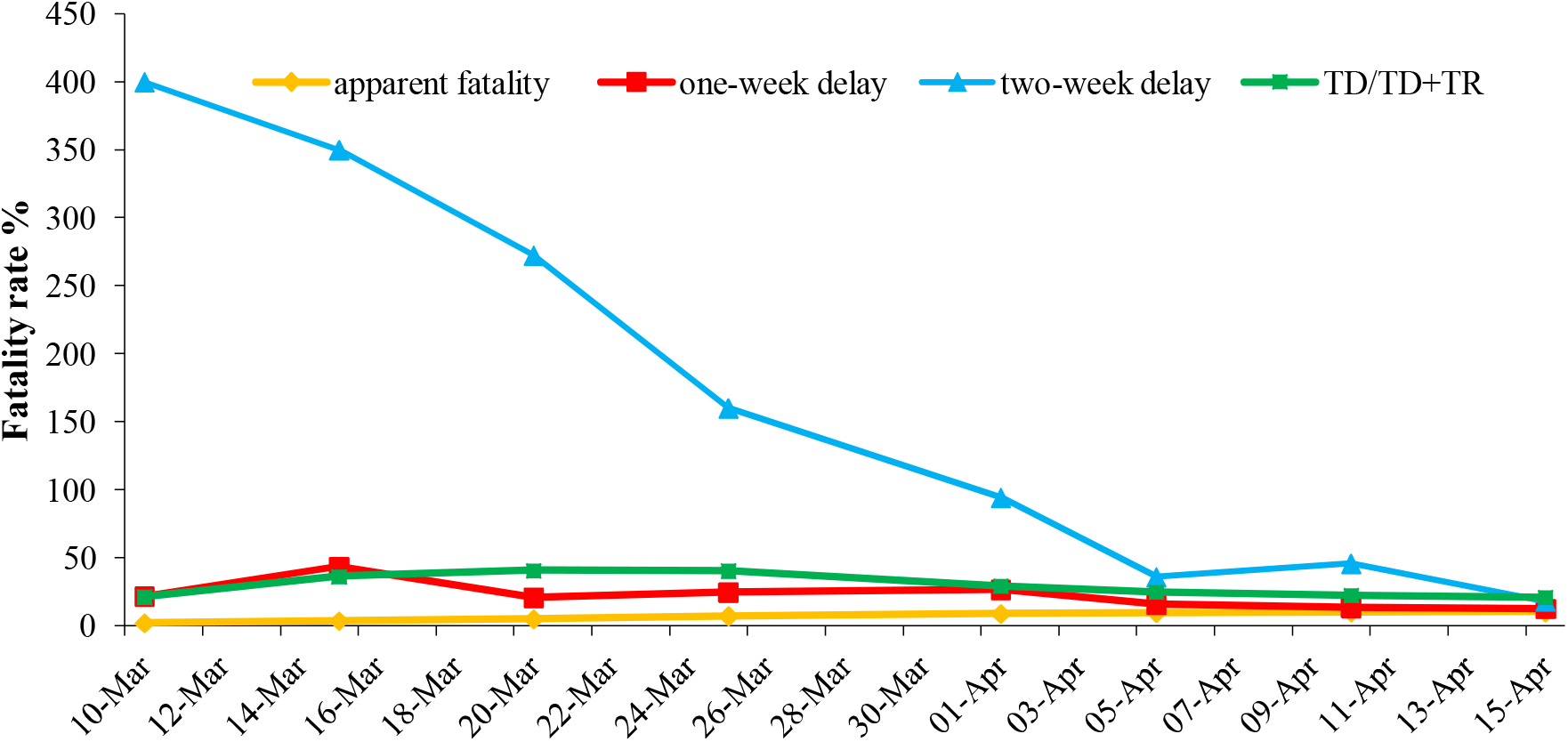
The fatality rate calculated based on four scenarios for Spain (data up to April 15^th^)

## Data Availability

All of used data are available online.

## Declarations

### Availability of data and materials

Not applicable

### Authors’ contributions

MY involved in the design and coordination of the study.AF helped in the design of the study MA involved in data collection and performed the statistical analysis. All authors read and approved the final manuscript.

### Consent for publication

Not applicable

## References

1. Battegay M, Kuehl R, Tschudin-Sutter S, Hirsch HH, Widmer AF, Neher RA. 2019-novel Coronavirus (2019-nCoV): estimating the case fatality rate–a word of caution. Swiss medical weekly. 2020;150.(0506)

2. Huang C, Wang Y, Li X, Ren L, Zhao J, Hu Y, et al. Clinical features of patients infected with 2019 novel coronavirus in Wuhan, China. The Lancet. 2020;395(10223):497–506.

3. Wang D, Hu B, Hu C, Zhu F, Liu X, Zhang J, et al. Clinical characteristics of 138 hospitalized patients with 2019 novel coronavirus–infected pneumonia in Wuhan, China. Jama. 2020.

4. Organization WH. Report of the who-china joint mission on coronavirus disease 2019 (covid-19). Available on-line: https://www.whoint/docs/default-source/coronaviruse/who-china-joint-mission-oncovid-19-final-reportpdf. 2020.

5. Baud D, Qi X, Nielsen-Saines K, Musso D, Pomar L, Favre G. Real estimates of mortality following COVID-19 infection. The Lancet Infectious Diseases. 2020.

6. Wang W, Tang J, Wei F. Updated understanding of the outbreak of 2019 novel coronavirus (2019‐nCoV) in Wuhan, China. Journal of medical virology. 2020;92(4):441–7.

